# Accuracy, Sensitivity, and Specificity of Different Tests to Detect Impaired Hand Function in Parkinson’s disease

**DOI:** 10.1101/2020.04.25.20079392

**Authors:** Cintia C. G. Alonso, Paulo Barbosa de Freitas, Raquel Pires, Dalton Lustosa de Oliveira, Sandra M. S. Ferreira de Freitas

## Abstract

**Introduction:** Parkinson’s Disease (PD) can affect hand function. To examine the diagnostic accuracy, sensitivity, and specificity of four traditionally used hand function tests in individuals with PD by using the receiver operator characteristic (ROC) analysis.

**Methods:** Eighty individuals (24 with PD and 56 healthy controls) performed the Jebsen-Taylor Hand Function Test (JTHF, with seven subtests), Nine-Hole Peg Test, and maximum power and pinch grip strength tests. The outcomes of the tests were compared between groups. The values of the area under the curve from the ROC analysis assessed the diagnostic accuracy, sensitivity, and specificity of the tests.

**Results:** Individuals with PD presented worst performance than controls in all tests, except the writing subtest of the JTHF and maximum power strength. Two subtests of the JTHF, the turning cards and moving large, heavy objects, showed the highest area under the curve in the ROC analysis. The Nine-Hole Peg Test was able to distinguish the PD stage and progression, while the simulated feeding of the JTHFT subtest showed a high area under the curve only for PD stage analysis.

**Conclusion:** Two dexterity tasks (turning cards and moving large, heavy objects) were highly discriminative of the hand function impairments in individuals with PD. The Nine-Hole Peg Test provides the most accurate identification of the PD stage and progression based on hand function impairment. Different dexterity tasks should be used depending on the aims of the evaluation, whether for diagnosis, monitoring, or classification of the PD.

## Introduction

The ability to perform manipulation tasks that are essential for daily living is affected in individuals with Parkinson’s disease, PD^1, 2^. In addition to the PD cardinal manifestations (tremor, bradykinesia, and rigidity), poor digits and hand dexterity and reduced strength have been responsible for the hand function impairments^3–7^. Therefore, the inclusion of one or more of hand function tests in the clinical examination may help the clinicians to provide a better quantitative estimate of the patient current sensorimotor condition and the disease progression, which are often based upon the Unified Parkinson’s Disease Rating Scale (UPDRS)^8^ and Hoehn and Yahr Scale^9^.

Among the standardized tests used to assess hand function in individuals with PD, the most common are the maximum isometric power and pinch grip strength^5, 10^, Digitized Archimedes Spiral Drawing^11^, and Nine-Hole Peg Test (9HPT)^4^. A recent study^6^ reported that the Jebsen-Taylor Hand Function Test (JTHFT), a widely used test for global hand function evaluation in post-stroke individuals^12, 13^, is adequate to assess the hand function of individuals with PD. It was observed that individuals with PD needed more time to complete the JTHFT than healthy controls, and the performance of their left hand was moderately correlated with the UPDRS score^8^. The timed dexterity tests can also detect small changes in hand function due to PD progression after two years^14^. However, these findings were based on the application of one of the hand function tests. It is unknown, for example, whether the performance in the JTHFT can better discriminate individuals with PD from controls than other simpler and less time-consuming hand function tests. Also, as the JTHFT is composed of seven subtests, it is not clear whether one or a few of those subtests could be enough to discriminate individuals with PD from controls.

Therefore, the current study aimed to test the ability of different hand function tests to discriminate individuals with PD from healthy controls. The diagnostic accuracy, sensitivity, and specificity of four hand function tests (described in methods section) were investigated in individuals with PD by the receiver operator characteristic (ROC) analysis. We hypothesized that although the group of individuals with PD would present worst performance in all tests when compared to a group of controls, a single test or a combination of few of them (we considered each subtest of the JTFHT as a separated test) could be more sensitive for hand function impairments in individuals with PD. The test that best discriminates individuals with PD from healthy controls could be routinely used for assessment in clinical settings and be selected as a primary or secondary outcome variable of clinical trials involving individuals with PD.

## Methods

### Participants

Eighty individuals, 24 individuals with PD (45 years and older) and 56 healthy controls, participated in this study. All participants were right-handed (confirmed by the Edinburg Handedness Inventory), had a normal or corrected vision, and tactile sensitivity preserved. Controls did not present any known or apparent musculoskeletal impairment in upper extremities or neurological disease. Individuals with PD were responsive to drug therapy and classified as stages I-III in the Hoehn and Yahr (H&Y) Scale. They were tested in their “on medication” status. Eight participants from the PD group were reassessed 18 months later. Participants gave informed consent as approved by the local human research ethics committee in accordance with the Declaration of Helsinki.

### Experimental Procedure

Four hand function tests, two assessing hand and digits dexterity (JTHFT and 9HPT), and two measuring maximum grip strength, were performed with the right and left hands. Half of the participants started with their right and a half with their left hand. First, participants performed the JTHFT (Model 8063, Sammons Preston, Bolingbrook, USA) and 9HPT (Rolyan Nine-Hole Peg Test, Model A8515, Sammons Preston) in an alternated order. Then, the power grip strength test (hydraulic hand dynamometer, Saehan, SH5005, Changwon, South Korea) followed by the pinch grip strength test (hydraulic pinch dynamometer, Saehan, SH5005, Changwon, South Korea) were performed. For all tests, participants remained seated in a regular chair in front of a table (except the strength tests) with adjusted height.

*JTHFT*. A single trial s Seven subtests simulating unimanual daily manipulation activities were performed in the following order as established by the original test protocol^13^: (1) writing; (2) turning cards; (3) moving small, common objects; (4) simulated feeding; (5) stacking checkers; (6) moving large, light objects; and (7) moving large, heavy objects. Each subtest was explained before its onset and participants were instructed to perform it as fast as they could. The writing task was performed according to the validated Brazilian version of the JTHFT^15^. A digital stopwatch was used to record the time of each subtest, and then these times were summed (JTHFT total). The time of each subtest and the sum of them were used for statistical analysis.

*9HPT*. A plastic rectangular console with a concave circular container in one side and nine small holes aligned in a 3-by-3 configuration on the other side was placed in front of the participant. Nine small cylindrical pegs (6.4mm in diameter and 32mm in length) were placed inside of the container. Participants were asked to pick a peg up from the container and place it into the hole, one by one until all holes were filled, and then, immediately, return the pegs, one by one, to their original container. Participants were instructed to perform this test as quickly as possible. A familiarization trial was performed before a valid one. The time to complete a single trial was recorded with a stopwatch.

*Strength Tests*. Participants remained seated with the arm aside the trunk, forearm pronated at 90° and horizontally oriented, and wrist slightly hyperextended. They hold a hand or pinch grip hydraulic dynamometer and squeezed them as hard as they could, keeping the force magnitude for about 4s and then relaxing. Three trials were performed using the whole hand for power grip strength and three trials using the index finger and thumb for pinch grip strength, with intervals of two minutes between trials to prevent fatigue. The highest values for power grip strength and pinch grip strength were selected and statistically analyzed.

### Statistical analyses

Despite being tested on both hands, we compared the hand of the PD onset, which was considered the most affected hand by the PD patients, with the non-dominant hand of the healthy control individuals. The non-dominant hand of the controls was selected because it showed the worst performance in all the tests when compared to the dominant hand.

The ability of the tests to discriminate individuals with PD and healthy controls was evaluated by the receiver operating characteristic (ROC) curve analyses. We calculated the area under the ROC curve (AUC), with values close to 1 suggesting high diagnostic accuracy of the test. The best cutoff point determined by the best combination of specificity and sensitivity using the Youden Index method^16^, the sensitivity, and specificity were presented for those variables with significant AUC. We also performed ROC curve analyses for combinations of variables with AUC>0.8. For all statistical tests, alpha values were set at 0.05. Statistical analyses were performed in IBM-SPSS (version 25).

## Results

Participants’ characteristics were similar between PD and control groups (Table 1). Most of the PD patients were classified as in stage I of H&Y (n=11), while the others were in stages II (n=9) or III (n=4). For 13 PD individuals, the symptoms started on the right side and for 11 participants, on the left side. The comparisons between the values of the outcomes of the tests performed with the dominant, right hand and non-dominant, left hand of the controls indicated a difference between hands for all tests (p<0.044), except for the subtest of the JTHFT turning cards. Controls spent more time to perform the tasks and were weaker with the non-dominant hand when compared to the dominant hand. For further analysis, values from the side of the PD onset considered the most affected side were compared with that from healthy individuals and the results are presented in Table 2.

**Table 1.** Characteristics and clinical results of PD and control groups.

|  | PD (N= 24) | Control (N=56) |
| --- | --- | --- |
| **Age (years)** | 66.3 ± 9.6 | 65.8 ± 9.7 |
| **Gender** | Male = 14<br>Female = 10 | Male = 24<br>Female = 32 |
| **Body height (cm)** | 165 ± 9.6 | 165 ± 11 |
| **Body mass (kg)** | 68 ± 16 | 76 ± 14 |
| **PD onset (years)** | 9.7 ± 4.3 | |
| **UPDRS** | 45 ± 14 | |
| **UPDRS (Part III)** | 21 ± 8 | |
| **Hoehn and Yahr Scale (Stages I-V)** | I = 11<br>II-III = 13 |  |
| **Onset side of PD** | Left= 11<br>Right= 13 |  |

*Note: Data are expressed as mean ± Standard Deviation, except gender, Hoehn and Yahr and onset side of PD. PD, Parkinson Disease; UPDRS, Unified Parkinson's Disease Rating Scale.*

The results of the ROC curve analyses revealed that the JTHFT total, power grip strength, and the time to perform the subtest writing of the JTHFT were not able to discriminate individuals with PD than controls. The results of the ROC curve analyses revealed that the performance in all other tasks discriminated individuals with PD and controls, with individuals with PD being slower and weaker than controls.

The AUC for the pinch grip strength was 0.72 (0.60-0.84, *p*=0.002). The maximum pinch grip strength can discriminate individuals with PD from controls with 87.5% of sensitivity and 51.8% of specificity (cutoff point=3.87kgf).

The AUCs for the JTHFT_noW_ and 9HPT were 0.88 (±95% confidence interval: 0.80-0.96, *p*<0.001) and 0.80 (0.70-0.91, *p*<0.001), respectively. The sum of the times to perform the six subtests of the JTHFT can discriminate individuals with PD from healthy controls with 91.7% of sensitivity and 73.2% of specificity (cutoff point=40.2s). The time to perform the 9HPT can discriminate individuals with PD from healthy controls with 66.7% of sensitivity and 82.1% of specificity (cutoff point=29.5s).

The ROC analysis carried out by the combination of the times to perform the JTHFT_noW_ and the 9HPT revealed an AUC of 0.87 (0.78-0.95, *p*<0.001). The sensitivity increased (83%, cutoff point=67.7s) compared to the 9HPT analyzed individually (66.7%), but it decreased compared to the sensitivity of only the JTHFT_noW_ (91.7%). The specificity in the combined performances was 75%, which is close to the JTHFT_noW_ alone (73.2%) and lower than the 9HPT alone (82.1%).

The ability of each JTHFT subtest (except the writing subtest) to differentiate individuals with PD and controls was also assessed with the ROC curve analyses. All subtests produced significant AUC values, ranging from 0.74 (stacking checkers) to 0.91 (moving large, heavy objects). The subtests turning cards and moving large, heavy objects presented the highest AUC values. The AUC for the turning cards subtest was 0.89 (0.82-0.96), with a sensitivity of 95.8% and specificity of 73.2% (cutoff point=5.92s). The AUC for the moving large, heavy objects was 0.91 (0.85-0.97), with a sensitivity of 91.7% and specificity of 73.2% (cutoff point=4.09s). The AUC for the moving large, light objects was 0.84 (0.75-0.94), with a sensitivity of 83.3% and specificity of 78.6% (cutoff point=4.34s). We combined the performance of the two subtests with the largest AUC (i.e., turning cards + moving large, heavy objects) and found an increase of less than 1% (AUC=0.92) when compared to the AUC of moving large, heavy objects alone. The combination of these two tests increased the sensitivity (100%), but the specificity was 75% (cutoff point=10.08s).

**Table 2.**
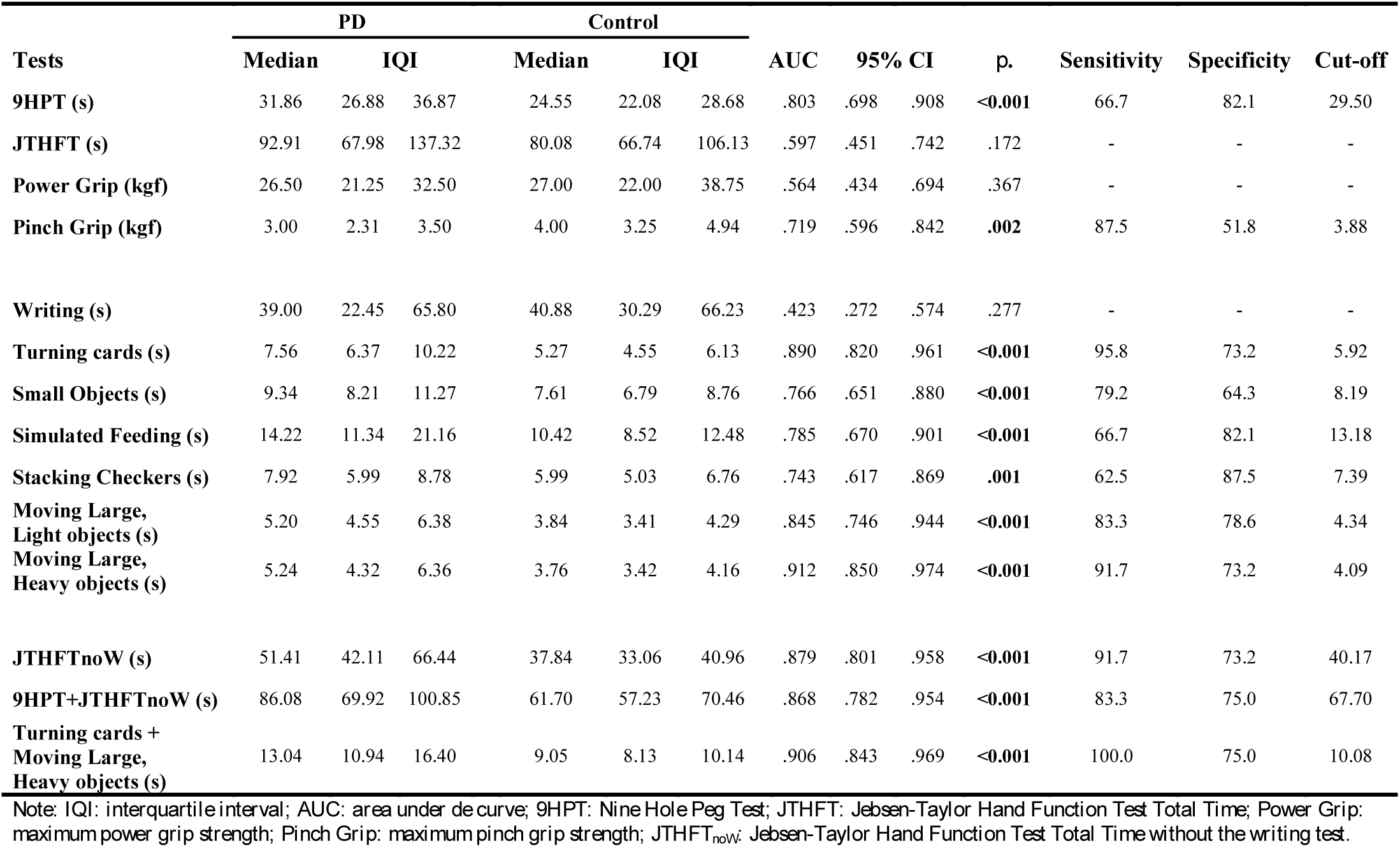
Outcomes of the hand function tests for PD and Control groups and the results of the statistical analyses.

### Stage discrimination

Based on the H&Y stages, the PD participants were divided into two groups (H&Y =1 and H&Y<1). Only two outcome variables were able to discriminate these groups: 9HPT and the simulated feeding JTHFT subtest. The AUC was 0.78 (0.60-0.97) for the 9HPT e 0.79 (0.59-0.98) for the subtest simulated feeding. The sensitivity for the 9HPT was 66.7% and the specificity was 75% (cutoff point=31.02s). For the simulated feeding subtest, the sensitivity was 58.3% and specificity was 100% (cutoff point=11.87s).

### Progression discrimination

Eight PD participants were reassessed after 18 months. Six participants had an increase in the UPDRS scores with an average of 7.3 points. Two participants classified as H&Y stage I were later classified as stage II. For this analysis, the comparison between assessments was performed for both more affected and less affected hands. Only the values of the 9HPT tests of the less affected hand increased after 18 months (p=0.038). The AUC for the 9HPT was 0.81 (0.58-1, p=0.04) and can discriminate the progression of PD after 18 months with a sensitivity of 75% and specificity of 87.5% (cutoff point=31.3s).

## Discussion

This study used different hand function tests in the same sample of individuals with PD to investigate which test or tests would provide the highest diagnostic accuracy in detecting hand function impairments in this population. Overall, individuals with PD showed worse performance in almost all hand function tests than controls, mainly in the ones requiring hand/digits dexterity. The performance in the tests requiring maximum force exertion was either unable to detect differences (i.e., power grip strength) or was moderately accurate in distinguishing individuals with PD and controls (i.e., pinch grip strength). The JTHFT without the subtest writing (JTHFT_noW_) and the 9HPT showed good diagnostic performance, with the JTHFT being more sensitive and the 9HPT more specific. The combination of the performance of the dexterity tests (JTHFT_noW_+9HPT) did not improve diagnostic accuracy. The two JTHFT subtests with the highest diagnostic accuracy were the turning cards and moving large, heavy objects. When they were combined, the diagnostic accuracy, sensitivity, and specificity increased.

Two hand function tests failed in discriminating individuals with PD and healthy controls: writing and maximum power grip strength. The finding that individuals with PD and controls presented a similar performance in writing contradicts other studies^11, 17–19^. In this study, participants were instructed to copy a sentence as fast and accurately as possible, and they may have prioritized accuracy in copying the sentence and not speed. Therefore, we conclude that a timing test requiring copying a sentence cannot differentiate individuals with PD and controls. However, we should acknowledge that writing a sentence and analyzing the words’ structure could provide valid information, as a large number of individuals with PD shown a smaller handwriting word size (micrographia) than healthy controls^20^.

Corroborating the results of previous studies^10, 21^, we found similar maximum power grip strength (AUC not significant) in PD and controls, suggesting that this test should not be indicated for detecting hand function impairment in this population. The values of maximum pinch grip strength, however, discriminated individuals with PD from controls, but with low sensitivity and specificity. The possible reason for the different results of strength tests is that small intrinsic muscles responsible for pinch grip are more affected by the PD^21^ than more proximal and bigger muscle groups (e.g., wrist flexors). Contrarily, pinch grip strength has not shown force deficits in individuals with PD^22^. Nevertheless, maximum hand and digits strength tests do not seem to be appropriate to detect impairment in individuals with PD in the H&Y Scale in stages between I to III.

Individuals with PD were slower than controls to perform the 9HPT (≈25%) as previously reported^4^, indicating that they have poorer digits dexterity than controls. Visuospatial information processing and high movement speed are crucial for fast performance in the 9HPT^23^. Movement slowness, one of the cardinal manifestations of PD, and deficits in the visuospatial information processing^23,24^ could explain the lower performance in the 9HPT by the individuals with PD. More important, however, is that the 9HPT is more accurate in discriminating (with high specificity and moderate sensitivity) the stage and progression of the disease than differentiating individuals with PD from controls. A previous study showed that the 9HPT had higher sensitivity and specificity to discriminate individuals with PD than the Grooved Pegboard Tests^25^, Purdue Pegboard, and Tapping tasks^26^.

The JTHFT has been described as a test for global hand function assessment^13^. The JTHFT subtests evaluate different hand function characteristics that have been used in post-stroke individuals^12, 27, 28^, individuals with diabetic peripheral neuropathy^28^, and, recently, in individuals with PD^6^. The fact that the total time of the JTHFT without the writing subtest shows an AUC close to 0.9 confirms the usefulness of this test in detecting hand function impairment in individuals with PD. Moreover, the performance in only two subtests was highly discriminative between PD and control groups: the turning cards and moving large, heavy objects. Both tests showed high AUC, with excellent sensitivity and moderate specificity ability, and when they were combined, a slight increase in sensitivity was observed but not in specificity. Specifically, the combination of these two tests produces a cutoff value that can discriminate all individuals with PD from healthy controls, but still, there are control adults who also perform the tests as slowly as some of the individuals with PD. Based on this latter result, the application of only these two JTHFT subtests would be recommended, saving time and effort of both evaluators and patients. The combination of the turning cards and moving large, heavy objects tests could be useful to detect hand function impairments in individuals with PD compared to healthy controls. On the other hand, the stage of the disease and the effects due to its progression were more accurately identified by the 9HPT, which assesses more fine motor dexterity. One question that remains is which test could be the best to assess the effects of motor rehabilitation.

This study is not exempt from limitations. The sample size could be considered relatively small for PD (N=24) and heterogeneous (e.g., H&Y stages I-III). Larger sample size would allow us to explore the ability of these tests to discriminate individuals with PD in more advanced stages of the H&Y Scale (e.g., II from III) and those in very early-stage PD (H&Y=I) from healthy controls. Also, a higher sample size would allow us to examine the ability of these tests to distinguish between individuals with PD tremor-dominant and those with postural instability and gait disability. However, even with the current sample, we found that a combination of two subtests requiring more speed is very sensitive in differentiating individuals with PD and age-matched healthy individuals as well as dexterity tests can identify the stage and progression of the disease. Future studies should explore the issues mentioned above and the possible use of those highly discriminative tests as auxiliary tools in the motor rehabilitation of PD.

## Conclusion

Individuals with PD present poorer hand dexterity and smaller maximum pinch grip strength than healthy controls, whereas they have similar speed when writing a sentence and maximum power grip strength. The tests with higher accuracy, sensitivity, and specificity in distinguishing individuals with PD and controls are two subtests of the JTHFT, i.e., turning cards and moving large, heavy objects. The combined performances in these two tests provides the highest discriminative accuracy. Hence, we conclude that the tasks involving more speed (less accuracy) are potentially able to accurately detect the impairments of the hand function in individuals with PD. Tasks involving more fine motor dexterity and speed might be used by clinicians and researchers to assess the progression of the hand function impairments in individuals with PD.

## Data Availability

Data will be available upon request.

## Acknowledgments

This study received support in part by the Coordenação de Aperfeiçoamento de Pessoal de Nível Superior – Brazil (CAPES) to Alonso – Finance Code 001. Freitas is grateful to CNPq/Brazil.

## References

1. Borrione P, Tranchita E, Sansone P, Parisi A. Effects of physical activity in Parkinson’s disease: A new tool for rehabilitation. World Journal of Methodology. 2014;4:133–143.

2. Mazzoni P, Shabbott B, Cortes JC. Motor control abnormalities in Parkinson’s disease. Cold Spring Harb. Perspect. Med. 2012;2:1–17.

3. Choi YI, Song CS, Chun BY. Activities of daily living and manual hand dexterity in persons with idiopathic parkinson disease. Journal of physical therapy science. 2017;29:457–460.

4. Earhart GM, Cavanaugh JT, Ellis T, Ford MP, Foreman KB, Dibble L. The 9-hole PEG test of upper extremity function: average values, test-retest reliability, and factors contributing to performance in people with Parkinson disease. J. Neurol. Phys. Ther. 2011;35:157–163.

5. Jones GR, Roland KP, Neubauer NA, Jakobi JM. Handgrip Strength Related to Long-Term Electromyography: Application for Assessing Functional Decline in Parkinson Disease. Arch. Phys. Med. Rehabil. 2017;98:347–352.

6. Mak MK, Lau ET, Tam VW, Woo CW, Yuen SK. Use of Jebsen Taylor Hand Function Test in evaluating the hand dexterity in people with Parkinson’s disease. J. Hand Ther. 2015;28:389–394.

7. Proud EL, Morris ME. Skilled hand dexterity in Parkinson’s disease: effects of adding a concurrent task. Arch. Phys. Med. Rehabil. 2010;91:794–799.

8. Movement Disorder Society Task Force on Rating Scales for Parkinson’s D. The Unified Parkinson’s Disease Rating Scale (UPDRS): status and recommendations. Mov. Disord. 2003;18:738–750.

9. Hoehn MM, Yahr MD. Parkinsonism: onset, progression and mortality. Neurology. 1967;17:427–442.

10. Villafane JH, Valdes K, Buraschi R, Martinelli M, Bissolotti L, Negrini S. Reliability of the Handgrip Strength Test in Elderly Subjects With Parkinson Disease. Hand. 2016;11:54–58.

11. Saunders-Pullman R, Derby C, Stanley K, et al. Validity of spiral analysis in early Parkinson’s disease. Mov. Disord. 2008;23:531–537.

12. Cunha BP, Freitas S, Freitas PB. Assessment of the Ipsilesional Hand Function in Stroke Survivors: The Effect of Lesion Side. J. Stroke Cerebrovasc. Dis. 2017;26:1615–1621.

13. Jebsen RH, Taylor N, Trieschmann RB, Trotter MJ, Howard LA. An objective and standardized test of hand function. Arch. Phys. Med. Rehabil. 1969;50:311–319.

14. Haaxma CA, Bloem BR, Overeem S, Borm GF, Horstink MW. Timed motor tests can detect subtle motor dysfunction in early Parkinson’s disease. Mov. Disord. 2010;25:1150–1156.

15. Ferreiro KN, Santos RL, Conforto AB. Psychometric properties of the portuguese version of the Jebsen-Taylor test for adults with mild hemiparesis. Rev. Bras. Fisioter. 2010;14:377–382.

16. Fluss R, Faraggi D, Reiser B. Estimation of the Youden Index and its associated cutoff point. Biometrical journal. Biometrische Zeitschrift. 2005;47:458–472.

17. San Luciano M, Wang C, Ortega RA, et al. Digitized Spiral Drawing: A Possible Biomarker for Early Parkinson’s Disease. PLoS One. 2016;11:1–11.

18. Stanley K, Hagenah J, Bruggemann N, et al. Digitized spiral analysis is a promising early motor marker for Parkinson Disease. Parkinsonism Relat. Disord. 2010;16:233–234.

19. Wang M, Wang B, Zou J, Nakamura M. A new quantitative evaluation method of spiral drawing for patients with Parkinson’s disease based on a polar coordinate system with varying origin. Physica A: Statistical Mechanics and its Applications. 2012;391:4377–4388.

20. Smits EJ, Tolonen AJ, Cluitmans L, van Gils M, Conway BA, Zietsma RC, et al. Standardized Handwriting to Assess Bradykinesia, Micrographia and Tremor in Parkinson’s Disease. PLoS ONE. 2014; 9(5): e97614.

21. Vaillancourt DE, Slifkin AB, Newell KM. Visual control of isometric force in Parkinson’s disease. Neuropsychologia. 2001;39:1410–1418.

22. Oliveira MA, Rodrigues AM, Caballero RMS, Petersen RD, Shim JK. Strength and isometric torque control in individuals with Parkinson’s disease. Exp. Brain Res. 2008;184:445–450.

23. Song CS. Relationship between visuo-perceptual function and manual dexterity in community-dwelling older adults. Journal of physical therapy science. 2015;27:1871–1874.

24. Raskin SA, Borod JC, Wasserstein J, Bodis-Wollner I, Coscia L, Yahr MD. Visuospatial orientation in Parkinson’s disease. Int. J. Neurosci. 1990;51:9–18.

25. Bohnen NI, Studenski SA, Constantine GM, Moore RY. Diagnostic performance of clinical motor and non-motor tests of Parkinson disease: a matched case-control study. Eur. J. Neurol. 2008;15:685–691.

26. Rudzinska M, Izworski A, Banaszkiewicz K, Bukowczan S, Marona M, Szczudlik A. Quantitative tremor measurement with the computerized analysis of spiral drawing. Neurol. Neurochir. Pol. 2007;41:510–516.

27. Jebsen RH, Griffith ER, Long EW, Fowler R. Function of “normal” hand in stroke patients. Arch. Phys. Med. Rehabil. 1971;52:170–174 passim.

28. Lima KCA, Borges LDS, Hatanaka E, Rolim LC, de Freitas PB. Grip force control and hand dexterity are impaired in individuals with diabetic peripheral neuropathy. Neurosci. Lett. 2017;659:54–59.

